# Kinesiophobia and physical activity: A systematic review and meta-analysis

**DOI:** 10.1101/2023.08.17.23294240

**Authors:** Miriam Goubran, Ata Farajzadeh, Ian M. Lahart, Martin Bilodeau, Matthieu P. Boisgontier

## Abstract

**Objective:** Physical activity contributes to the primary, secondary, and tertiary prevention of multiple diseases. However, in some patients, an excessive, irrational, and debilitating fear of movement (i.e., kinesiophobia) is thought to induce avoidance behaviors, contributing to decreased engagement in physical activity. The aim of this study was to examine whether kinesiophobia is negatively associated with physical activity in several health conditions and what factors may influence this relationship.

**Methods:** Five databases were searched for studies including both a measure of kinesiophobia and physical activity. Two reviewers screened articles for inclusion, assessed risk of bias, and extracted data from each study. Pearson product-moment correlations were pooled from eligible studies using the generic inverse pooling and random effects method to examine the relationship between kinesiophobia and physical activity.

**Results:** Seventy-four studies were included in the systematic review and 63 studies (83 estimates, 12,278 participants) in the main meta-analysis. Results showed a small-to-moderate negative correlation between kinesiophobia and physical activity (r = −0.19; 95% confidence interval: −0.26 to −0.13; I^2^ = 85.5%; p < 0.0001). Funnel plot analysis showed evidence of publication bias, but p-curve analysis suggested that our results could not be caused by selective reporting. A subgroup meta-analysis showed that the correlation was statistically significant in patients with cardiac, rheumatologic, neurologic, or pulmonary conditions, but not in patients with chronic or acute pain.

**Conclusion:** Our results suggest that higher levels of kinesiophobia are associated with lower levels of physical activity in several health conditions that are not necessarily painful.

**Impact:** Kinesiophobia should be dissociated from pain and considered in relation to specific health conditions when implementing exercise therapy. Kinesiophobia may have prognostic implications in patients for whom physical activity contributes to prevent recurrence or worsening of their condition.

## INTRODUCTION

Seven decades ago, the seminal work of Morris et al. (1953)^1^ showed that conductors on London double-decker buses, who were responsible for checking tickets, assisting passengers with luggage, and supervising the loading and unloading of passengers, had a lower incidence and less severe coronary heart disease than bus drivers. Since then, the scientific literature demonstrating the health benefits of physical activity has grown exponentially and expanded to include multiple health conditions^2^. These benefits include reduced risk of disability, disease, and mortality^2,3^. Specifically, higher levels of physical activity have been shown to contribute to a reduced risk of cardiovascular disease^4^, obesity^5^, depression^6^, hypertension^7^, cancer^8^, and dementia^9^. Yet, one in four adults worldwide does not meet the recommendations for physical activity^10^. Physical activity also plays an important role in secondary and tertiary prevention by reducing the impact, slowing the progression, and preventing the recurrence of multiple health conditions, including cardiovascular disease^11,12^, osteoarthritis^13^, stroke^14,15^, and cancer^16^.

Several factors may explain physical inactivity^17^, including environmental, interpersonal, and intrapersonal factors^18^. Environmental factors include lack of access, weather conditions, and safety concerns^19^. Interpersonal factors include family responsibilities, lack of support, and lack of a gym partner^20^. Intrapersonal factors include gender^21^, age^22^, cognitive function^23,24^, and socioeconomic circumstances^25^. Another intrapersonal factor of interest is kinesiophobia, which can be defined as an excessive, irrational, and debilitating fear of movement and activity resulting from a sense of vulnerability to pain, injury, or a medical condition^26^. Kinesiophobia is typically measured using self-administered questionnaires, such as the Tampa Scale of Kinesiophobia (TSK)^27,28^, which assesses an individual’s belief that physical activity can lead to injury or pain and that the severity of their medical condition is underestimated. While kinesiophobia is often observed in the context of pain or a clinical condition, its presence in otherwise healthy adults is also possible^29^ due to the irrational nature of this phobic condition. The irrational fear that characterizes kinesiophobia is likely to influence the desires and impulses for movement and rest^30^, as well as affective determinants of physical activity in general^31^.

The relationship between kinesiophobia and physical activity can be explained by theories suggesting that the perception of a cue related to physical activity automatically activates the concept of physical activity as well as the unpleasant (or pleasant) affective memories associated with this concept^32–35^. This activation results in an impulse that favors the tendency to avoid (or approach) physical activity^36^. Thus, negative affective associations are likely to hinder physical activity. Accordingly, an aversive fear of pain, injury, or aggravation of a medical condition that has been associated with the concept of movement may result in the development of automatic avoidance behaviors that contribute to the maintenance and exacerbation of this fear, and ultimately lead to a phobic state (i.e., kinesiophobia) that diminishes the ability to engage in regular physical activity.

Previous systematic syntheses of the literature on this topic include a meta-analysis^37^ and two systematic reviews^38,39^. The main results of these reviews suggest that exercise interventions may reduce kinesiophobia in individuals with back pain. While back pain is one condition that may contribute to kinesiophobia, it is not the only one. The relationship between physical activity and kinesiophobia should be investigated in other conditions such as cardiac, neurological, and rheumatologic conditions.

The main objective of this study was to systematically review and meta-analyze the direct relationship between kinesiophobia and physical activity. We hypothesized that levels of kinesiophobia would be negatively associated with levels of physical activity. In addition, we examined the moderating effect of health status, physical activity measurement instruments (i.e., accelerometers, pedometers, questionnaires), physical activity outcomes (e.g., total physical activity, moderate or vigorous physical activity, steps per day), and kinesiophobia measurement instruments. Finally, because kinesiophobia and physical activity can vary with age, sex, and pain^40,41^, we explored the influence of these factors on the association between kinesiophobia and physical activity.

## METHODS

### Search Strategy

This review was reported according to the Preferred Reporting Items for Systematic Reviews and Meta-Analyses (PRISMA) guidelines^42^. Potential studies were identified by searching the MEDLINE (via PubMed), PsychInfo, CINAHL, EMBASE, and SPORTDiscus databases. In October 2023, two reviewers (MG and AF) searched for all available records using the following combination of keywords in the title or abstract of the article: (“kinesiophobia” OR “fear avoidance” OR “fear of movement” OR “movement phobia” OR “movement fear”) AND (“physical activity” OR “exercise” OR “walking”). In PsychInfo the limits “clinical trial”, “quantitative study”, “peer-reviewed journal”, “English”, and “human” were used. In PubMed the limits “clinical trial”, “observational study”, “RCT”, “English” were used. In SPORTDiscus the limits “peer-reviewed”, “English”, “academic journal”, and “article” were used. In CINAHL the limits “peer-reviewed”, “English”, “research article”, “journal article”, and “humans” were used. To reduce literature bias^43,44^, this systematic review was pre-registered in PROSPERO^45^.

### Eligibility Criteria and Study Selection

#### Inclusion Criteria

To be included in this systematic review, articles had to be published in a peer-reviewed journal, be written in English, report original data collected from human participants, include at least one self-reported measure of kinesiophobia and one measure of physical activity, and formally test the association between these two variables, be it a univariate or multivariate test. The physical activity measure could be derived from a self-reported measure of the level of physical activity or from a device (e.g., accelerometer, pedometer) worn while participants are engaged in their normal daily activities.

#### Exclusion Criteria

Studies were excluded if they were published as a book chapter, study protocol, conference abstract, or were based on laboratory-based measures of physical fitness (e.g., maximal muscle force, 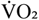 max) and not on a measure of physical activity.

#### Study Selection

Article screening was performed in Covidence systematic review software (Veritas Health Innovation, Melbourne, Australia; www.covidence.org), a web-based collaborative software platform that streamlines the production of systematic reviews. After removing duplicates, titles and abstracts were independently reviewed by two reviewers (MG, AF) according to the inclusion and exclusion criteria using a systematic 5-step process. If there was any doubt at any step, the full text was further reviewed. Step 1: Articles not written in English were excluded. Step 2: Articles that did not report original empirical data were excluded (e.g., reviews, meta-analyses, commentaries, technical reports, case studies). Step 3: Articles that did not involve human participants were excluded. Step 4: Articles that did not assess both kinesiophobia and physical activity were excluded. Step 5: Articles that did not formally test the association between kinesiophobia and physical activity were excluded. In addition, we performed reference screening and forward citation tracking on the articles remaining after step 5. Any disagreements between the two reviewers were resolved by consensus among three reviewers (MG, AF, MPB).

### Data Extraction

Data extracted from selected articles included first author’s name, article title, publication year, digital object identifier (DOI), number of participants, number of men and women, age range, mean age, mean weight, mean height, mean body mass index, health status, mean pain intensity, type of kinesiophobia measure, level of kinesiophobia, type of physical activity measure, type of physical activity outcome, level of physical activity (continuous or categorical), as well as statistical estimates and significance of the association between kinesiophobia and physical activity.

### Methodological Quality and Risk of Bias Assessment

The risk of bias of the studies included in the systematic review was estimated using the National Institutes of Health (NIH) Quality Assessment Tool for Observational Cohort and Cross-Sectional Studies^46^, the Transparent Reporting of Evaluations with Non-Randomized Designs (TREND) reporting checklist^47^, and the Consolidated Standards of Reporting Trials (CONSORT) reporting checklist for randomized trials^48^. All scores were normalized to a 0-10 scale to make them comparable across assessment instruments (Table 1).

**Table 1.**
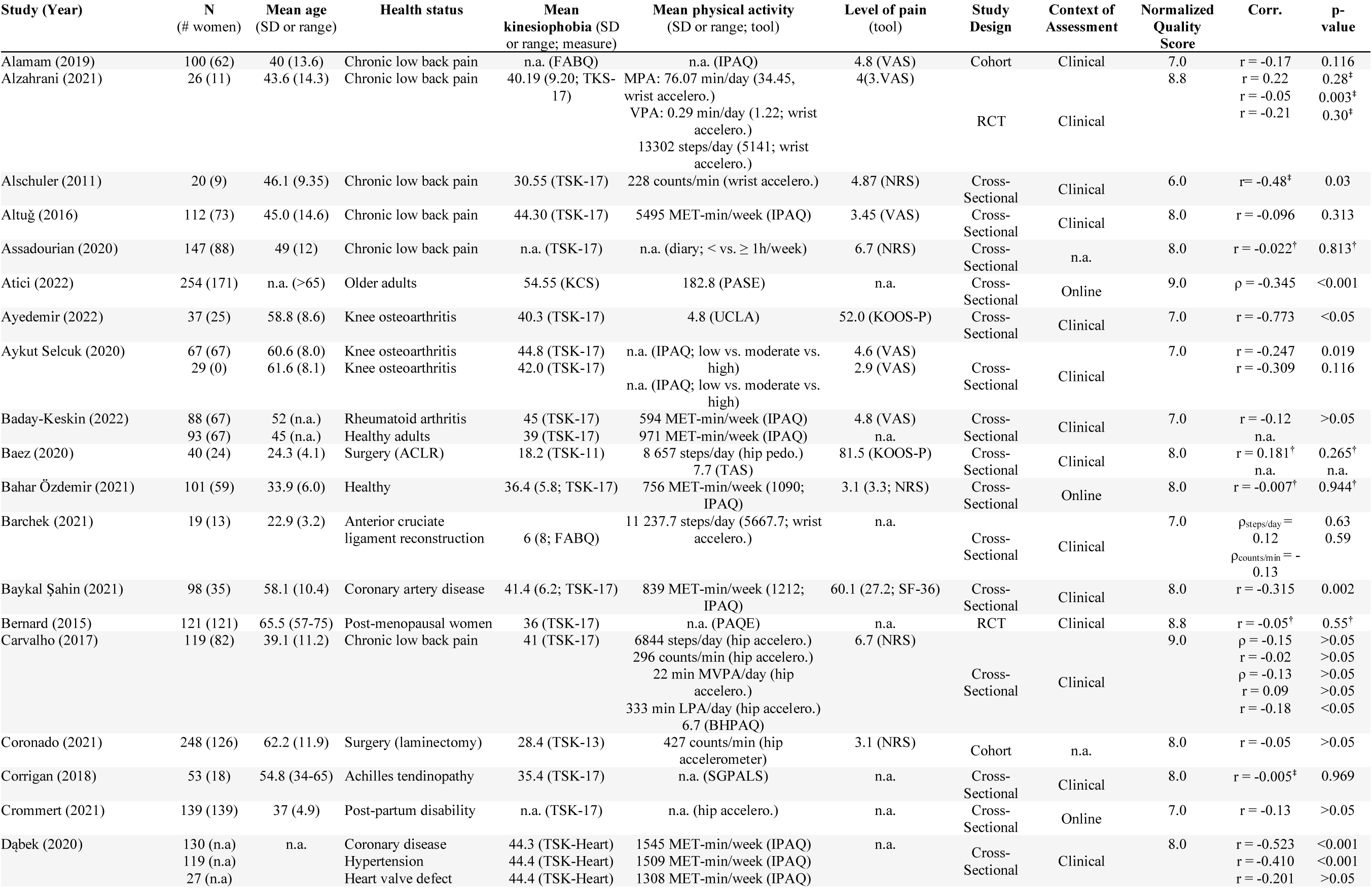
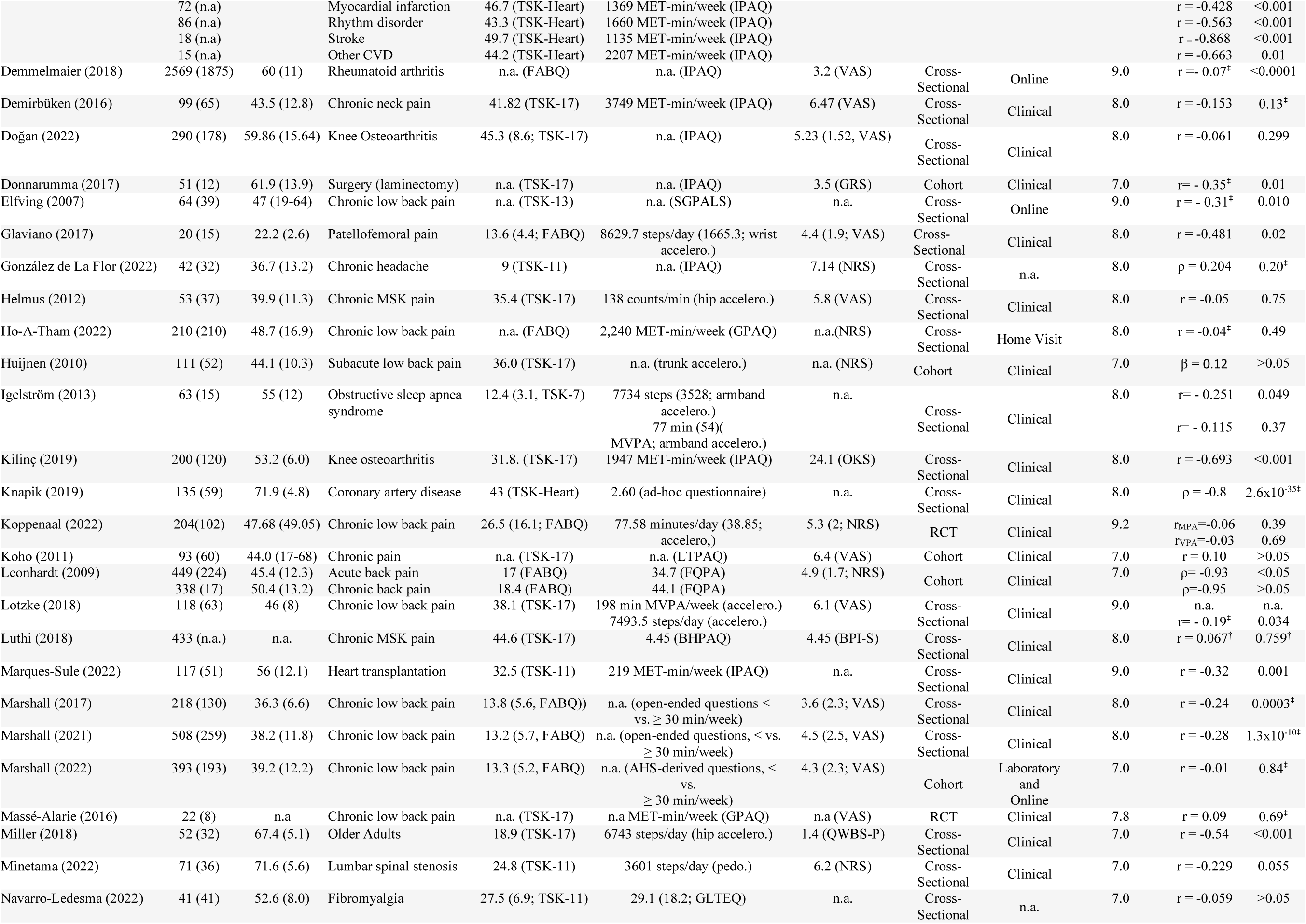
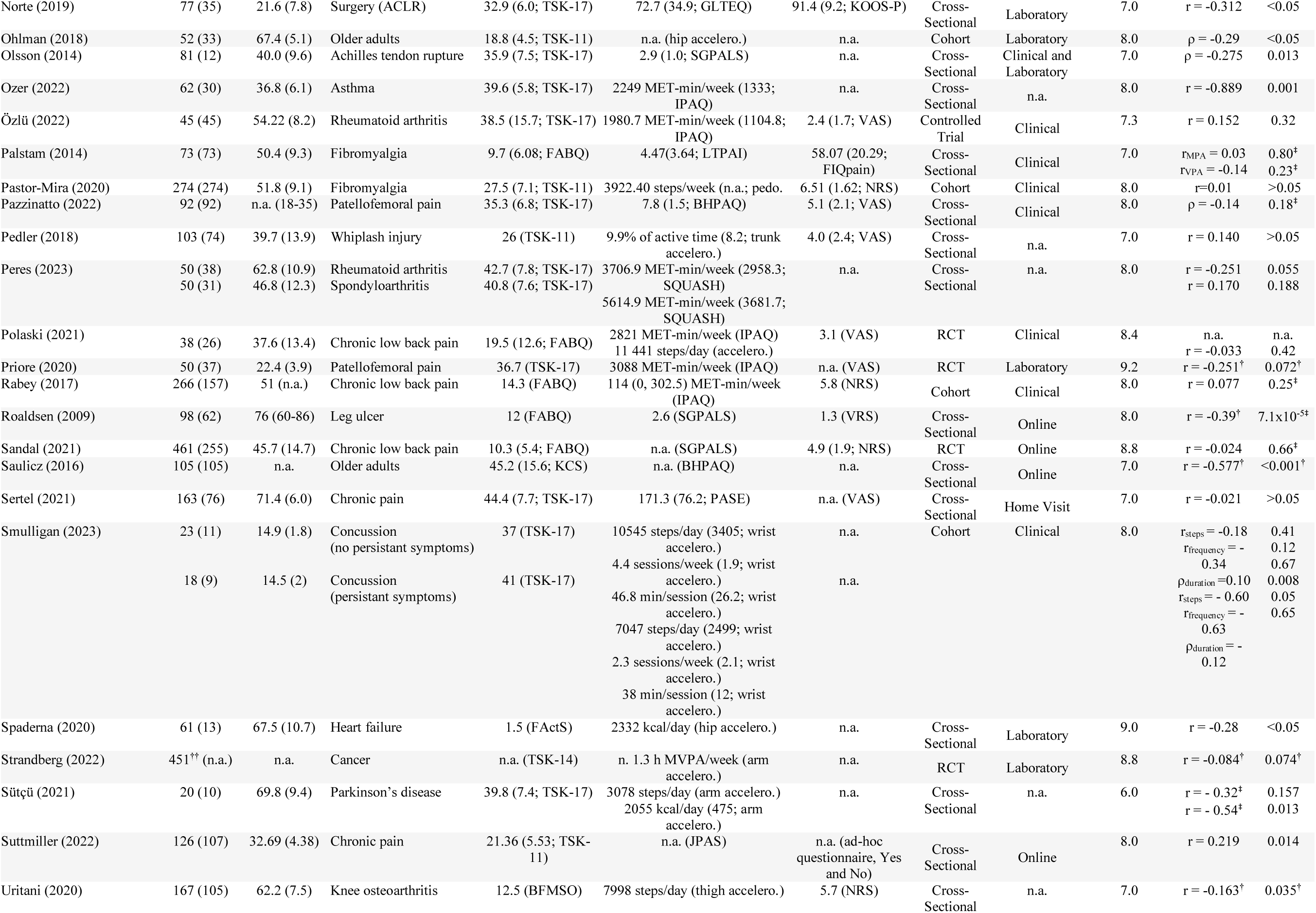
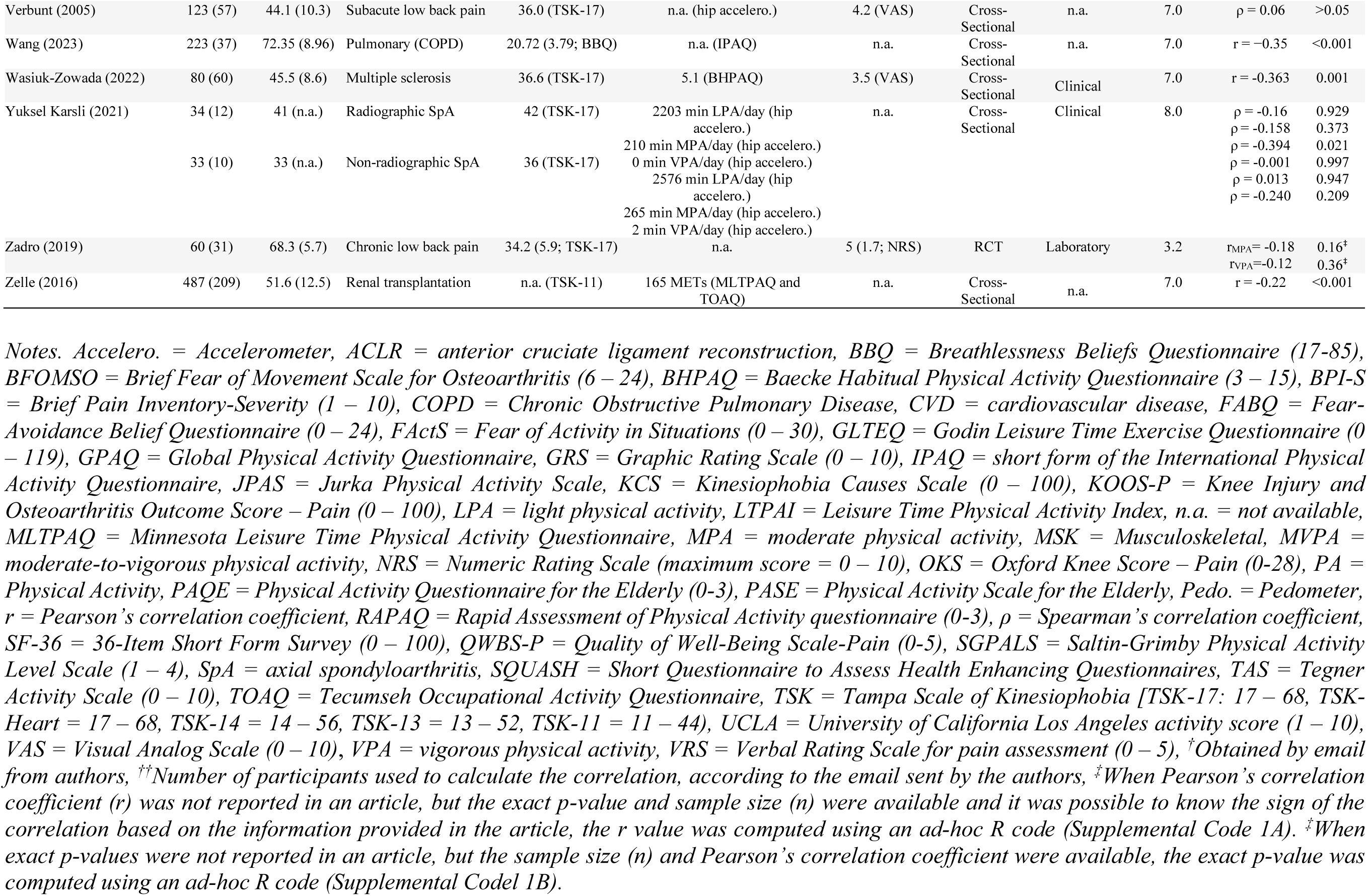
Sample characteristics of studies included in the systematic review.

### Meta-Analysis

All analyses were performed in R Studio integrated development environment (IDE) (2023.06.1+524, “Mountain Hydrangea” release) for R software environment^49^ using the {meta}^50^ and {metafor}^51,52^ R packages^53^.

#### Main Meta-Analysis

We pooled Pearson product-moment correlations from eligible studies to examine the relationship between kinesiophobia and physical activity. Correlations were pooled using the generic inverse pooling method via the ‘metacor’ function in the {meta} R package^50^. This function automatically performs a necessary Fisher’s z-transformation on the original, untransformed correlations prior to pooling. The ‘metacor’ function also reconverts the pooled association back to its original form for ease of interpretation. Correlation estimates were nested within studies using the ‘cluster’ argument to account for the dependencies between these estimates, resulting in a three-level meta-analysis (level 1: participants, level 2: correlation estimates, level 3: studies). The distribution of variance across levels was assessed using the multilevel version of I^2^ ^54^. The performance of the 2-level and 3-level meta-analyses was assessed and compared using the {metafor} R package^51,52^. We anticipated considerable between-study heterogeneity, and therefore used a random-effects model to pool correlations. The restricted maximum likelihood (RML) estimator^55^ was used to calculate the heterogeneity variance Tau^2^. In addition to Tau^2^, to quantify between-study heterogeneity, we report the I^2^ statistic, which provides the percentage of variability in the correlations that is not caused by sampling error^56^. The I^2^ statistic was interpreted as follows: 0-40%, may not be important; 30-60%, may represent moderate heterogeneity; 50-90%, may represent substantial heterogeneity; and 75-100%, may represent considerable heterogeneity. To reduce the risk of false positives, we used a Knapp-Hartung adjustment^57^ to calculate the confidence interval around the pooled association. We also report the prediction interval, which provides a range within which we can expect the associations of future studies to fall based on the current evidence. The pooled correlation was interpreted using Cohen’s conventions^58^: r ≈ −0.10, small negative correlation; r ≈ −0.30, moderate negative correlation; r ≈ −0.50, large negative correlation.

#### Publication Bias Assessment

Publication bias was assessed using a funnel plot, which is a scatter plot of the studies’ effect size expressed as the Fisher’s z transformed correlation on the x-axis against a measure of their standard error (which is indicative of precision of the study’s effect size) on the y-axis. When there is no publication bias, the data points in a funnel plot should form a roughly symmetrical, upside-down funnel. Studies in the top part of the plot, which have lower standard errors, are expected to lie closely together, and not far away from the pooled effect size. In the lower part of the plot, studies have higher standard errors, the funnel “opens up”, and effect sizes are expected to scatter more heavily to the left and right of the pooled effect. Egger’s regression^59^ can be used to formally test funnel plot’s asymmetry. However, since there is no direct function to conduct Egger’s test for 3-level models, we calculated it by using the standard errors of the effect size estimates as a predictor in the meta-regression^60^.

P-curve analysis^61^ was conducted to assess whether the distribution of the statistically significant results was consistent with what would be expected if only true effects were present. When the null hypothesis is true (i.e., there is no true effect), p-values are assumed to follow a uniform distribution: highly significant effects (e.g., p = 0.01) are as likely as barely significant effects (e.g., p = 0.049). However, when the null hypothesis is false (i.e., there is a true effect in our data), p-values are assumed to follow a right-skewed distribution: highly significant effects are more likely than barely significant effects. A left-skewed distribution would suggest that some studies used statistical tests to find significant results in ways that may not be reproducible or generalizable (i.e., p-hacking).

#### Secondary Meta-Analysis

A secondary meta-analysis was conducted using the same approach, but based on Spearman’s rho values, to further test the relationship between kinesiophobia and physical activity.

#### Subgroup analyses and meta-regressions

Subgroup analyses were conducted to examine the differences in correlations between studies including participants with different health conditions and using different types of physical activity measures (i.e., device-based versus self-reported), physical activity measurement instruments (i.e., type of questionnaires, type of devices), physical activity outcomes, and kinesiophobia measures. Exploratory meta-regressions were conducted to examine if the average age of participants, the proportion of women, and pain in a study predicted the reported correlation between kinesiophobia and physical activity. Pain was normalized to a 0-100 scale to make the data comparable across pain scales. A sensitivity analysis was conducted to examine whether the quality of the studies affected the results.

## RESULTS

### Literature Search

The primary search identified 3,015 potentially relevant articles from the five databases (Figure 1), including 912 duplicates. Of the 2,103 articles screened, disagreement occurred in 210 cases (10%), all of which were resolved by consensus. All articles remained after step 1 as they were all written in English. 1,133 articles were excluded in step 2 because they were irrelevant (n = 710) or did not report original data (n = 423). No articles were excluded in step 3 because they all involved human participants. Eight hundred and fifty-two articles were excluded in step 4 because they did not assess kinesiophobia (n = 117) or physical activity (n = 735). Seventy-seven articles were initially excluded at step 5 because they did not formally test the correlation between kinesiophobia and physical activity or did not report the estimate of this correlation. However, the corresponding authors of these articles were contacted by email to request the Pearson correlation estimate of this association and the sample size used to calculate it. Nineteen authors replied to our email: Eight authors provided raw data for 10 studies^62–71^ and 11 authors provided the Pearson’s correlation estimate^29,72–81^. In addition, the Pearson’s correlation estimate of two articles were calculated based on information reported in the article^82,83^. This process reduced the number of studies excluded at step 5 to 54, resulting in a total of 64 articles included from the databases. Using reference screening and forward citation tracking, the authors identified 27 studies that assessed both physical activity and kinesiophobia, of which 8 reported an estimate of their relationship^84–91^ and 19 did not^92–110^. The corresponding authors of these 19 studies were asked by email to provide this estimate or their data. Two authors sent the estimate^104,107^. Seventeen emails remained unanswered^92–103,105,106,108–110^.

**Figure 1.**
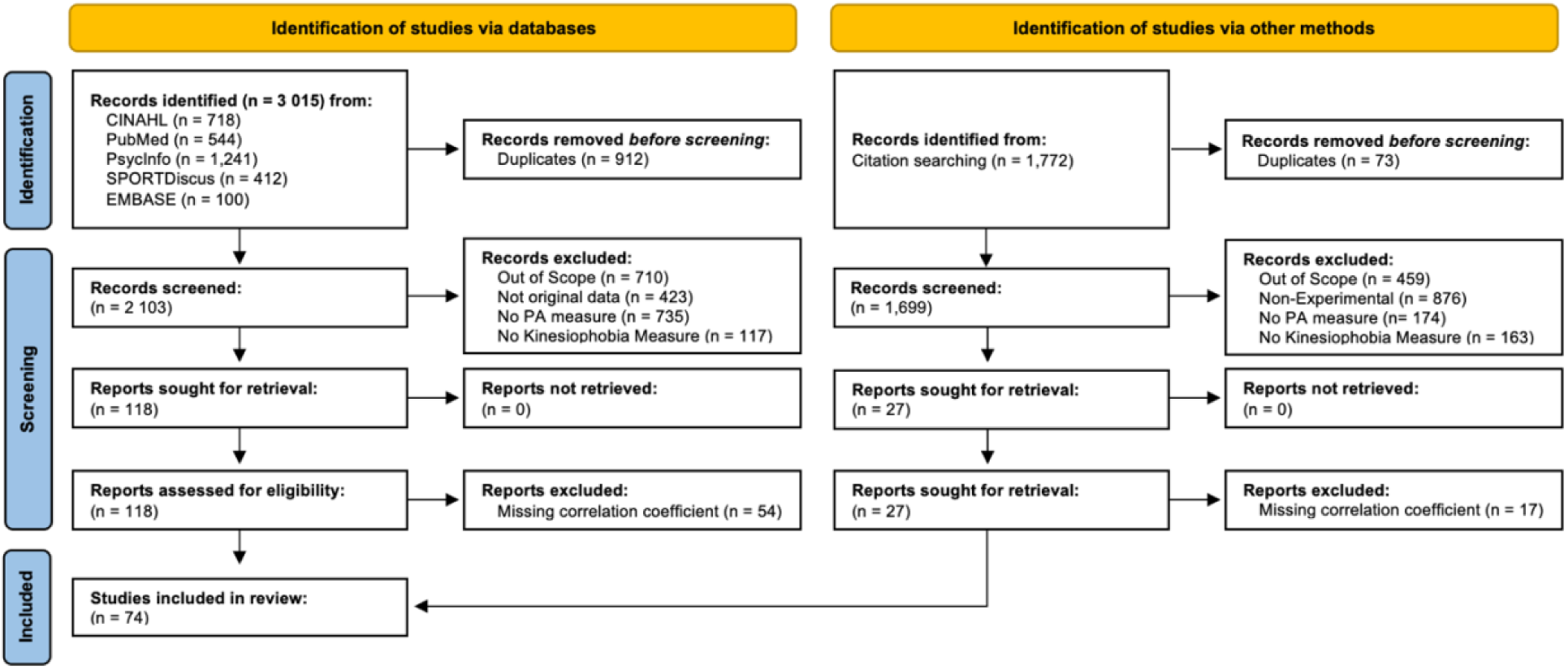
PRISMA 2020 flow diagram

### Descriptive Results

#### Participants

The 74 articles identified by the systematic review included a total of 13388 participants aged 11 to 85 years, including 7308 women, 4729 men, and 1351 participants whose gender and sex was not reported. The studies investigated populations with pain (n = 37)^62,64–71,73,76–81,83,88,89,111–128^ cardiac conditions (n = 6)^86,87,104,129–131^, surgery (n = 8)^63,131–137^, arthritis (n = 10)^75,82,84,85,91,138–142^, neurologic conditions (n = 3)^90,143,144^, pulmonary conditions (n = 3)^145–147^, cancer (n = 1)^74^, women health conditions (n = 2)^72,148^, as well as healthy adults (n = 6)^29,85,107,149–151^ (Table 1).

#### Kinesiophobia

In 54 of the 74 studies, kinesiophobia was assessed using the 17-item TSK (TSK-17; n = 38)^29,62,64,65,68,72,73,77,84,85,90,91,111–114,117–120,122–125,130,133–135,138–144,146,148,150^ shorter versions of the TSK [(TSK-11^152^; n = 10)^63,81,88,116,121,122,128,131,136,151^, (TSK-14; n = 1)^74^, (TSK-13^153^; n = 2)^115,137^, (TSK-7; n = 1)^145^], or its adaptation for patients with coronary artery disease (TSK-Heart^154^; n = 2)^86,87^. The TSK is a questionnaire that assesses the belief that movements can lead to (re)injury, pain, or aggravation of an underlying and serious medical condition^28^. Each item is rated on a Likert scale ranging from 1 (strongly disagree) to 4 (strongly agree). On the TSK-17, a score of 37 is used to distinguish between low (≤ 37) and high (> 37) levels of kinesiophobia^27^. On the TSK-13, scores inferior to 23 are considered sub-clinical^155^. The other measures that were used are the Fear-Avoidance Belief Questionnaire^156^ (FABQ; n = 15)^66,67,69–71,76,78–80,82,83,104,126,127,132^, Kinesiophobia Causes Scale^157^ (KCS; n = 2)^107,149^, the Fear of Activities in Situations scale (FActS; n = 1)^129^, the Brief Fear of Movement Scale for Osteoarthritis^158^ (BFMSO; n = 1)^75^, and Breathlessness Beliefs Questionnaire (BBQ; n = 1)^147^.

Sixty-four studies reported mean levels of kinesiophobia (Table 1). The studies based on the TSK-17 or TSK-Heart (mean range: 17 to 68) reporting the highest levels of kinesiophobia were those involving participants with a cardiovascular condition (41.4 to 49.7), followed by studies testing participants with arthritis (31.8 to 45.27) or chronic pain (30.5 to 44.6). Levels of kinesiophobia were lower in participants with a neurological (36.6 to 41), pulmonary (20.7 to 39.6), women health (36), or surgical condition (32.9 to 35.9), and in healthy adults (18.9 to 39.0).

#### Physical Activity

Fifty-one studies assessed physical activity using a self-reported measure (Table 1). Most of these questionnaire-based studies used the short form of the International Physical Activity Questionnaire (IPAQ-SF; n = 20)^29,73,76,78,79,82,85,86,112,114,116,130,131,133,138–141,146,147^ which consists of 6 items assessing time spent in light (i.e., walking), moderate (e.g., carrying light loads, cycling at moderate speed, doubles tennis), and vigorous physical activity (e.g., digging, fast cycling, heavy lifting, aerobics) over the last 7 days^159^. Other questionnaires were used to assess physical activity, such as the Baecke Habitual Physical Activity Questionnaire^160^ (BHPAQ; n = 5)^64,89,90,107,113^, Saltin-Grimby Physical Activity Level Scale^161^ (SGPALS; n = 5)^80,104,115,125,135^, Godin-Shephard Leisure-Time Exercise Questionnaire^162^ (GLTEQ; n = 2)^121,134^, Minnesota Leisure Time Physical Activity Questionnaire^163^ (MLTPAQ; n = 1)^136^, Physical Activity Scale for the Elderly^164^ (PASE; n = 2)^123,149^, Physical Activity Questionnaire for the Elderly^164^ (PAQE; n = 1)^72^, Short Questionnaire to Assess Health Enhancing Physical Activity^165^ (SQUASH; n = 1)^142^, the Tegner Assessment Scale^166^ (TAS; n = 1)^63^, University of California Los Angeles (UCLA) activity score^167^ (n = 1)^84^, Leisure Time Physical Activity Index^168^ (LTPAI; n = 1)^67^, Global Physical Activity Questionnaire^169^ (GPAQ; n = 2)^77,83^, Freiburger Questionnaire on Physical Activity^170^ (FQPA; n = 1)^127^, Jurka Physical Activity Scale^171^ (JPAS; n = 1)^81^, Rapid Assessment of Physical Activity questionnaire^172^ (RAPAQ; n = 1)^68^, Tecumseh Occupational Activity Questionnaire^173^ (TOAQ; n = 1)^136^, and Australian Health Survey-derived questions (AHS; n = 1)^71^.

Physical activity was also assessed with devices such as accelerometers measuring accelerations in 3 dimensions (n = 23)^65,66,74,75,78,91,110,113,117,118,120,122,124,126,129,132,137,143–145,148,150,151^ and pedometers measuring the number of steps (n = 3)^63,88,128^ (Table 1). In most studies, the device was worn at the hip (n = 10)^63,91,113,117,124,129,137,148,150,151^. Other positions included wrist (n = 5)^65,111,126,132,143^, arm (n = 3)^74,144,145^, trunk (n = 2)^118,122^, and thigh (n = 1)^75^, with five studies not reporting where the device was worn^66,78,88,120,128^. Most studies that employed accelerometer-based measures used the ActiGraph (Actigraph, LLC, Pensacola, FL, USA) GT3X+ (n = 4)^120,137,150,151^, wGT3X-BT (n = 2)^138,148^ or GT9X Link (n = 2)^78,132^. The other accelerometers were the RT3 (Stayhealthy Inc., Monrovia, CA, USA; n = 3)^117,118,124^, the SenseWear Pro3 Armband (BodyMedia, Pittsburgh, PA, USA; n = 3)^74,144,146^, the Activity Sensory Move II (movisens GmbH, Karlsruhe, Germany; n = 1)^129^, the LifeShirt (Vivometrics, Inc., Ventura, CA, USA; n = 1)^122^, the ActiWatch (Mini Mitter Co., Inc., Bend, OR, USA; n = 1)^111^, AX3 (Axtivity, Newcastle upon Tyne, UK; n = 1)^65^, FitBit (FitBit Inc., San Francisco, CA) Charge HR (n = 1)^126^, Charge 3 (n = 1)^143^, and the Activ8 (2M Engineering, North Brabant, Netherland, n = 1)^66^. The type of accelerometer was not reported in one study^113^. The pedometers were the Digi-Walker SW-200 (New Lifestyles Inc., Lees Summit, MO, USA; n = 1)^63^, the Active Style Pro HJA-350IT (Omron Heathcare, Kyoto, Japan; n = 1)^88^ and Yamax Power-Walker EX-510 3D (Pedometer Express, Minnesota, USA; n = 1)^128^. These devices were worn for 5 days (n = 1)^111^, 6 days (n = 1^129^), 7 days (n = 16)^63,65,75,88,91,113,117,118,120,124,132,137,144,148,150,151^, or 14 days (n = 1)^126^. The remaining 7 studies did not specify the number of days the device was worn^66,74,78,122,128,143,145^. All studies provided the accelerometer or pedometer on the day kinesiophobia was assessed (n = 18)^63,65,66,75,78,88,89,111,113,120,122,126,132,137,143,148,150^. The remaining studies did not specify whether kinesiophobia was measured the day the device was provide or the last day of physical activity assessment (n = 7)^74,118,124,128,144,146,151^.

To assess physical activity, the studies used the following outcomes: Score from a questionnaire (e.g., TAS, PAQE, BHPAQ, SGPALS, LTPAQ, n = 24)^63,64,67,68,72,80,81,84,87,89,90,104,107,113,115,119,121,123,125,127,134,137,149^, MET-min/week (n = 23)^31,73,76,77–79,83,85,86,112,114,116,130,131,133,136,138–142,146,147^, steps per day (n = 14)^63,65,75,78,88,113,120,126,128,132,143–145,150^, hours per day or week (n = 12)^62,65,69–71,74,91,113,120,138,143,145^, counts per minute (n = 4)^111,113,117,137^, kilocalories per day (n = 2)^129,144^, or percentage of active time (n = 1)^122^. Nine studies used multiple physical activity outcomes^63,65,78,91,113,120,143–145^.

#### Association Between Physical Activity and Kinesiophobia

Among the 74 articles included in the systematic review, 42 reported correlation coefficients of the association between physical activity and kinesiophobia. Specifically, 32 articles reported at least one Pearson’s r correlation coefficient and 12 articles reported at least one Spearman’s rho^87,89,91,113,116,124,127,132,135,143,149,151^. When a correlation coefficient was not reported, but the exact p-value (or t value) and sample size were available and it was possible to know the sign of the correlation, which was the case for 7 studies^83,111,115,120,125,133,144^, the Pearson’s r estimate was computed using an ad-hoc R code (Supplemental Code 1A). For the studies that reported a relative p-value < 0.001 instead of an exact p-value, we used a p-value of 0.0009 to estimate an approximate r value^82^.

Through email correspondence with the authors, we obtained 23 additional Pearson’s r estimates^29,62–81,104,107^. In total, 83 Pearson’s r estimates from 63 studies and 21 Spearman’s rho estimates from 12 studies were used in the meta-analysis (Table 1). The remaining study did not report a correlation coefficient and was therefore not included in the meta-analysis^117^. This study reported a non-statistically significant positive association between physical activity and kinesiophobia based on a standardized beta coefficient.

#### Pain

Mean pain intensity at rest was reported in 45 out of the 74 articles included in the systematic review. Most studies used the Visual Analog Scale^174^ (VAS; n = 21)^65,69–71,76,78,82,85,89,90,112,114,117,119,120,122,124,126,138,139,141^ or the Numeric Rating Scale^175^ (NRS; n =15)^29,62,66,68,75,79,80,83,88,111,113,116,127,128,137^. Other studies used the Knee Injury Osteoarthritis Outcome Score pain subscale^176^ (KOOS-P; n = 3)^63,84,134^, Brief Pain Inventory^177^ (n = 1)^64^, Oxford Knee Score^178^ (OKS; n = 1)^140^, the Quality of Well-Being Scale – Self-administered Pain Scale^179^ (QWBS-P; n = 1)^150^, the Short Form 36 bodily pain^180^ (SF-36; n = 1)^130^, the Graphic Rating Scale^181^ (GRS; n = 1)^133^, Fibromyalgia Impact Questionnaire-Pain^182^ (FIQ-Pain, n = 1)^67^ and the Verbal Rating Scale^183^ (VRS; n = 1)^104^. In the meta-analysis, scores that were not on a 0-100 scale in the initial measure were scaled to that range.

### Meta-Analysis

#### Main Meta-Analysis

Our main meta-analysis of 63 studies, 83 Pearson’s r correlation estimates, and 12278 participants revealed a statistically significant small-to-moderate negative correlation between kinesiophobia and physical activity (r = −0.19; 95% confidence interval [95CI]: −0.26 to −0.13; p < 0.0001) (Table 2; Figure 2). However, we observed substantial-to-considerable between-study statistical heterogeneity (Tau^2^ = 0.06, 95CI: 0.02 to 0.09; I^2^ = 85.5%, 95CI: 82.6 to 87.9%), and the prediction interval ranged from r = −0.605 to 0.300, indicating that a moderate positive correlation cannot be ruled out for future studies.

**Figure 2.**
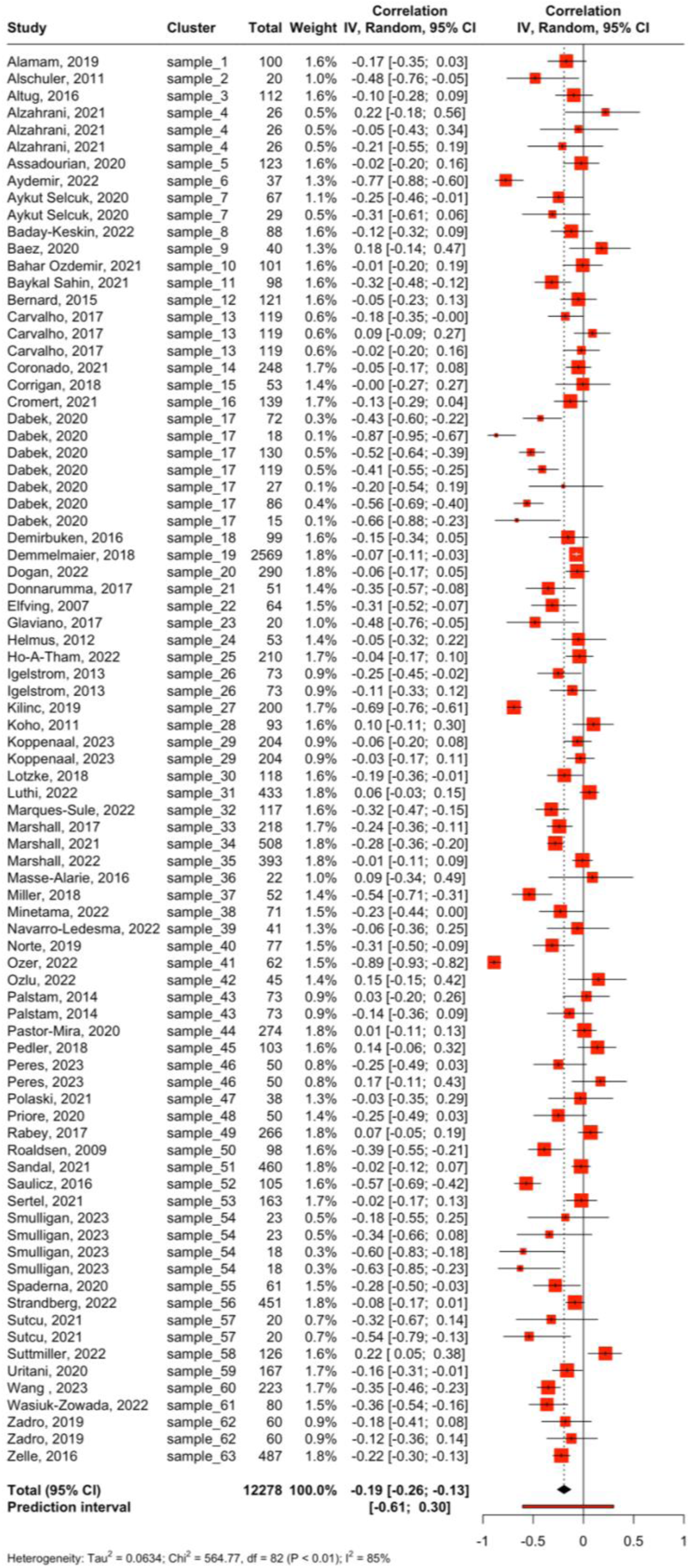
Main meta-analysis: Correlation between kinesiophobia and physical activity *Notes:* 95% CI = 95% confidence interval, IV = Inverse variance method, Random = Random effects method.

**Table 2.**
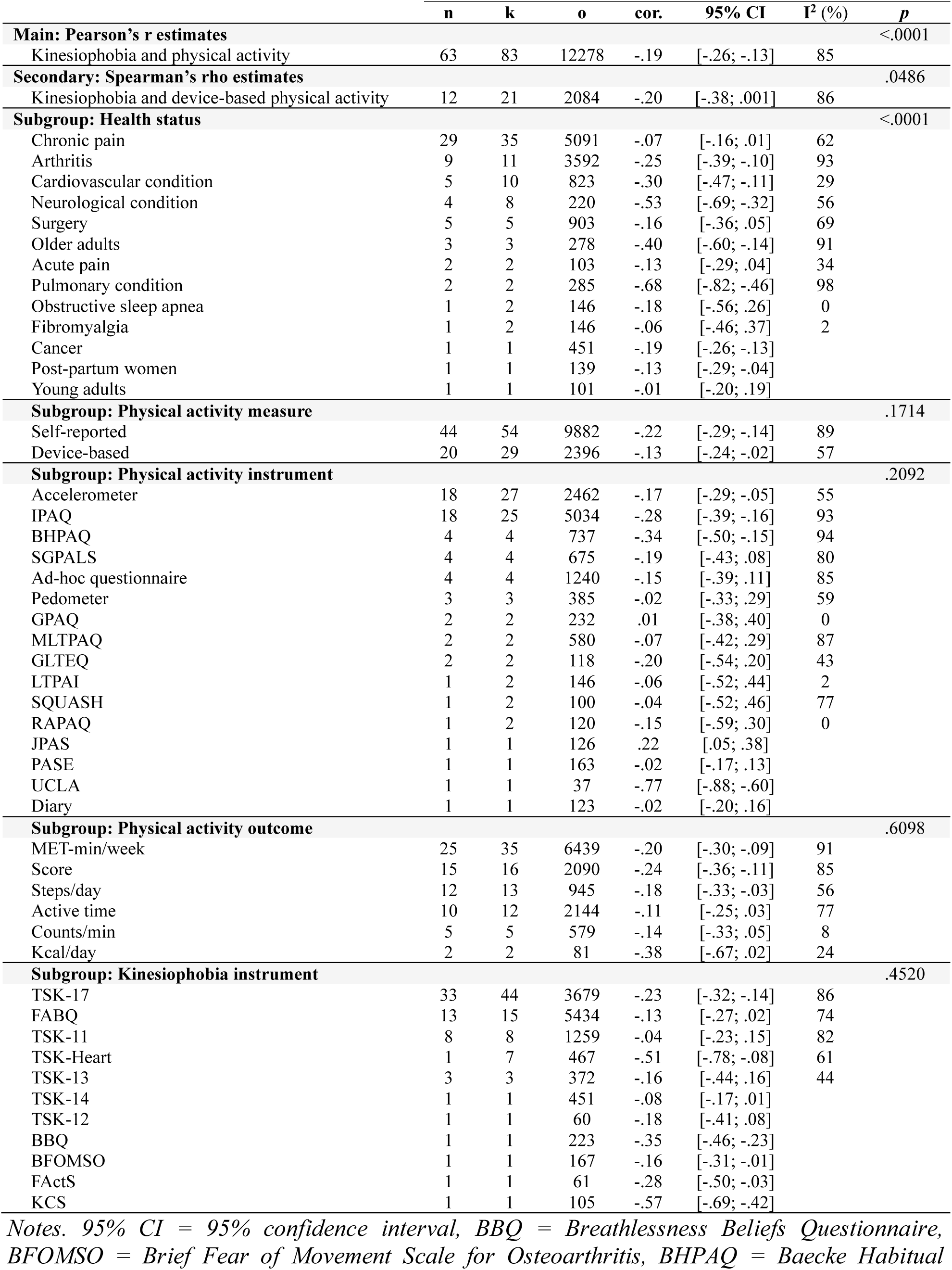
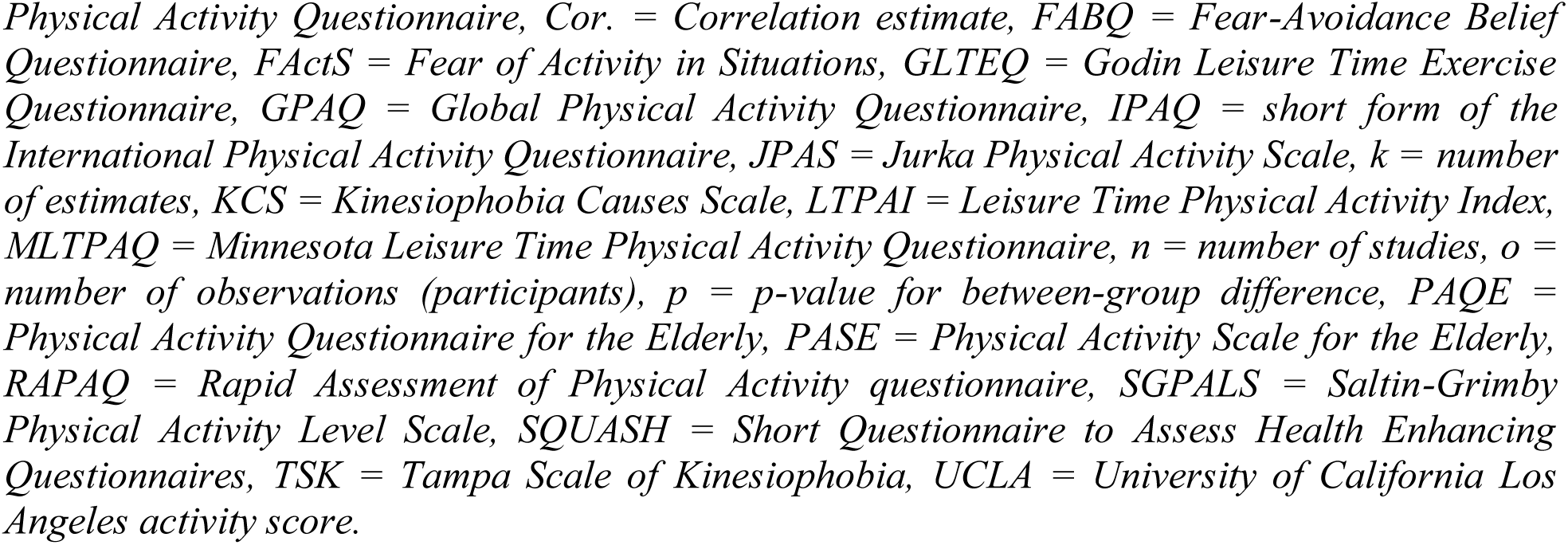
Results of the main, secondary, and subgroup meta-analyses.

|  | **n** | **k** | **o** | **cor.** | **95% CI** | **I<sup>2</sup> (%)** | **p** |
| --- | --- | --- | --- | --- | --- | --- | --- |
| **Main: Pearson's r estimates** |  |  |  |  |  |  | <.0001 |
| Kinesiophobia and physical activity | 63 | 83 | 12278 | -.19 | [-.26; -.13] | 85 |  |
| **Secondary: Spearman's rho estimates** |  |  |  |  |  |  | .0486 |
| Kinesiophobia and device-based physical activity | 12 | 21 | 2084 | -.20 | [-.38; .001] | 86 |  |
| **Subgroup: Health status** |  |  |  |  |  |  | <.0001 |
| Chronic pain | 29 | 35 | 5091 | -.07 | [-.16; .01] | 62 |  |
| Arthritis | 9 | 11 | 3592 | -.25 | [-.39; -.10] | 93 |  |
| Cardiovascular condition | 5 | 10 | 823 | -.30 | [-.47; -.11] | 29 |  |
| Neurological condition | 4 | 8 | 220 | -.53 | [-.69; -.32] | 56 |  |
| Surgery | 5 | 5 | 903 | -.16 | [-.36; .05] | 69 |  |
| Older adults | 3 | 3 | 278 | -.40 | [-.60; -.14] | 91 |  |
| Acute pain | 2 | 2 | 103 | -.13 | [-.29; .04] | 34 |  |
| Pulmonary condition | 2 | 2 | 285 | -.68 | [-.82; -.46] | 98 |  |
| Obstructive sleep apnea | 1 | 2 | 146 | -.18 | [-.56; .26] | 0 |  |
| Fibromyalgia | 1 | 2 | 146 | -.06 | [-.46; .37] | 2 |  |
| Cancer | 1 | 1 | 451 | -.19 | [-.26; -.13] |  |  |
| Post-partum women | 1 | 1 | 139 | -.13 | [-.29; -.04] |  |  |
| Young adults | 1 | 1 | 101 | -.01 | [-.20; .19] |  |  |
| **Subgroup: Physical activity measure** |  |  |  |  |  |  | .1714 |
| Self-reported | 44 | 54 | 9882 | -.22 | [-.29; -.14] | 89 |  |
| Device-based | 20 | 29 | 2396 | -.13 | [-.24; -.02] | 57 |  |
| **Subgroup: Physical activity instrument** |  |  |  |  |  |  | .2092 |
| Accelerometer | 18 | 27 | 2462 | -.17 | [-.29; -.05] | 55 |  |
| IPAQ | 18 | 25 | 5034 | -.28 | [-.39; -.16] | 93 |  |
| BHPAQ | 4 | 4 | 737 | -.34 | [-.50; -.15] | 94 |  |
| SGPALS | 4 | 4 | 675 | -.19 | [-.43; .08] | 80 |  |
| Ad-hoc questionnaire | 4 | 4 | 1240 | -.15 | [-.39; .11] | 85 |  |
| Pedometer | 3 | 3 | 385 | -.02 | [-.33; .29] | 59 |  |
| GPAQ | 2 | 2 | 232 | .01 | [-.38; .40] | 0 |  |
| MLTPAQ | 2 | 2 | 580 | -.07 | [-.42; .29] | 87 |  |
| GLTEQ | 2 | 2 | 118 | -.20 | [-.54; .20] | 43 |  |
| LTPAI | 1 | 2 | 146 | -.06 | [-.52; .44] | 2 |  |
| SQUASH | 1 | 2 | 100 | -.04 | [-.52; .46] | 77 |  |
| RAPAQ | 1 | 2 | 120 | -.15 | [-.59; .30] | 0 |  |
| JPAS | 1 | 1 | 126 | .22 | [.05; .38] |  |  |
| PASE | 1 | 1 | 163 | -.02 | [-.17; .13] |  |  |
| UCLA | 1 | 1 | 37 | -.77 | [-.88; -.60] |  |  |
| Diary | 1 | 1 | 123 | -.02 | [-.20; .16] |  |  |
| **Subgroup: Physical activity outcome** |  |  |  |  |  |  | .6098 |
| MET-min/week | 25 | 35 | 6439 | -.20 | [-.30; -.09] | 91 |  |
| Score | 15 | 16 | 2090 | -.24 | [-.36; -.11] | 85 |  |
| Steps/day | 12 | 13 | 945 | -.18 | [-.33; -.03] | 56 |  |
| Active time | 10 | 12 | 2144 | -.11 | [-.25; .03] | 77 |  |
| Counts/min | 5 | 5 | 579 | -.14 | [-.33; .05] | 8 |  |
| Kcal/day | 2 | 2 | 81 | -.38 | [-.67; .02] | 24 |  |
| **Subgroup: Kinesiophobia instrument** |  |  |  |  |  |  | .4520 |
| TSK-17 | 33 | 44 | 3679 | -.23 | [-.32; -.14] | 86 |  |
| FABQ | 13 | 15 | 5434 | -.13 | [-.27; .02] | 74 |  |
| TSK-11 | 8 | 8 | 1259 | -.04 | [-.23; .15] | 82 |  |
| TSK-Heart | 1 | 7 | 467 | -.51 | [-.78; -.08] | 61 |  |
| TSK-13 | 3 | 3 | 372 | -.16 | [-.44; .16] | 44 |  |
| TSK-14 | 1 | 1 | 451 | -.08 | [-.17; .01] |  |  |
| TSK-12 | 1 | 1 | 60 | -.18 | [-.41; .08] |  |  |
| BBQ | 1 | 1 | 223 | -.35 | [-.46; -.23] |  |  |
| BFOMSO | 1 | 1 | 167 | -.16 | [-.31; -.01] |  |  |
| FActS | 1 | 1 | 61 | -.28 | [-.50; -.03] |  |  |
| KCS | 1 | 1 | 105 | -.57 | [-.69; -.42] |  |  |

Notes. 95% CI = 95% confidence interval, BBQ = Breathlessness Beliefs Questionnaire, BFOMSO = Brief Fear of Movement Scale for Osteoarthritis, BHPAQ = Baecke Habitual
*Physical Activity Questionnaire, Cor. = Correlation estimate, FABQ = Fear-Avoidance Belief Questionnaire, FActS = Fear of Activity in Situations, GLTEQ = Godin Leisure Time Exercise Questionnaire, GPAQ = Global Physical Activity Questionnaire, IPAQ = short form of the International Physical Activity Questionnaire, JPAS = Jurka Physical Activity Scale, k = number of estimates, KCS = Kinesiophobia Causes Scale, LTPAI = Leisure Time Physical Activity Index, MLTPAQ = Minnesota Leisure Time Physical Activity Questionnaire, n = number of studies, o = number of observations (participants), p = p-value for between-group difference, PAQE = Physical Activity Questionnaire for the Elderly, PASE = Physical Activity Scale for the Elderly, RAPAQ = Rapid Assessment of Physical Activity questionnaire, SGPALS = Saltin-Grimby Physical Activity Level Scale, SQUASH = Short Questionnaire to Assess Health Enhancing Questionnaires, TSK = Tampa Scale of Kinesiophobia, UCLA = University of California Los Angeles activity score.*

The sampling error variance on level 1 and the value of I^2^ on level 2, i.e., the amount of heterogeneity variance within studies, were small (10.3% and 8.2%, respectively). The largest share of heterogeneity variance was from level 3, with between-study heterogeneity making up 81.5% of the total variation in our data (Supplemental Figure 1). Overall, this indicates that there is considerable between-study heterogeneity, and less than one tenth of the variance can be explained by differences within studies.

The 3-level model showed a better fit than the 2-level model with lower Akaike’s information criterion (AIC) (28.4 vs. 39.0) and Bayesian information criterion (BIC) (35.6 vs. 43.8), indicating better performance. These lower AIC and BIC are consistent with the significant likelihood ratio test (LRT) comparing the two models (χ^2^ = 12.67, p = 0.0004). Therefore, although the 3-level model introduces an additional parameter, this added complexity has improved our estimate of the pooled effect.

#### Publication bias assessment

Egger’s regression test using the standard errors of the effect size estimates as a predictor in the meta-regression showed that the coefficient of the standard error was significant (b = −1.497, 95CI: −2.618 to −0.3754, p = 0.0095), suggesting that the data in the funnel plot was asymmetrical (Figure 3A). This asymmetry may be explained by publication bias, but also by other potential causes, such as different study procedures and between-study heterogeneity^184^, which was substantial-to-considerable here.

**Figure 3.**
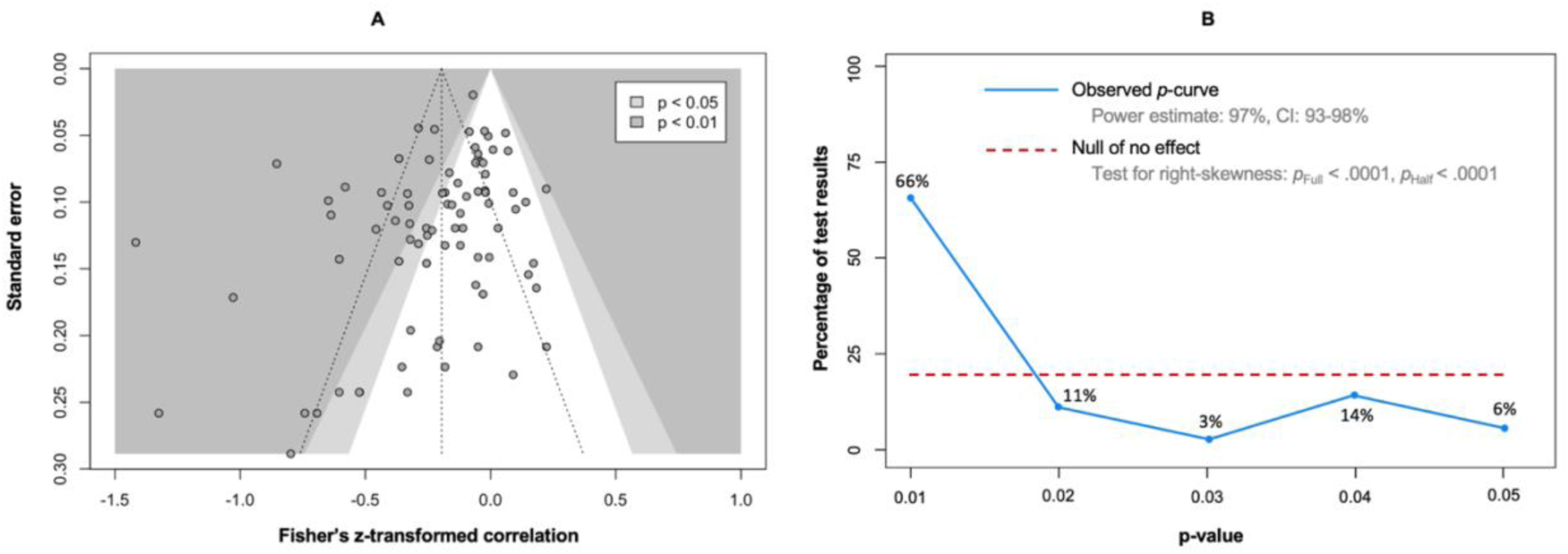
Publication bias assessment. Contour-enhanced funnel plot of the main meta-analysis (A). The vertical dashed line represents the average effect size. The two other dashed lines represent the idealized funnel-shape that studies are expected to follow. P-curve analysis (B). The blue line indicates the distribution of the analyzed p-values. The red dotted line illustrates a uniform distribution of the p-values, indicating the absence of a true effect.

The 83 Pearson’s r correlation estimates were provided to the p-curve analysis. The observed p-curve included 35 statistically significant results (p < 0.05), 27 of which were highly significant (p < 0.025), and was visually right-skewed (Figure 3B). The other results were excluded because they had a p > 0.05. The p-value of the right-skewness test was < 0.001 for both the half curve (curve of p-values ≤ 0.025) and the full curve (curve of p-values < 0.05), confirming that the p-curve was right-skewed and suggesting that the effect of our meta-analysis is true, i.e., that the effect we estimated is not an artifact caused by selective reporting (e.g., p-hacking) in the literature^185^. In addition, the statistical power of the studies that were included in the p-curve analysis was 97% (90CI: 93 to 98%), suggesting that approximately 90% of the significant results are expected to be replicable.

#### Secondary Meta-Analyses

Results of the secondary meta-analysis of 12 studies, 21 Spearman’s rho correlation estimates, and 2084 participants was consistent with the main meta-analysis as it showed a statistically significant small-to-moderate negative correlation between kinesiophobia and physical activity (r = −0.20; 95CI: −0.38 to −0.01; p = 0.049) (Table 2; Supplemental Figure 2). However, we observed substantial-to-considerable between-study statistical heterogeneity (Tau^2^ = 0.10, 95CI: 0.04 to 0.28; I^2^ = 86.3%) and the prediction interval ranged from r = −0.710 to 0.445, indicating that a moderate positive correlation cannot be ruled out for future studies.

#### Subgroup Meta-Analyses

The test of subgroup differences between health status was conducted on studies comprising people with chronic (k = 35) or acute pain (k = 2), arthritis (k = 11), a cardiovascular condition (k = 10), a neurological condition (k = 8), surgery (k = 5), older age (k = 3), obstructive sleep apnea (k = 2), a pulmonary condition (k = 2), fibromyalgia (k = 2), cancer (k = 1), as well as in post-partum women (k = 1) and healthy young adults (k = 1) (Table 2; Supplemental Figure 3). We found a statistical moderating effect of health status (p = 0.0014). The relationship between kinesiophobia and physical activity was statistically significant only in studies that included participants with cardiac condition (r = −0.30; 95CI: −0.47 to −0.11), arthritis (r = −0.25; 95CI: −0.39 to −0.10), a neurologic condition (r = −0.53; 95CI: −0.69 to −0.32), a pulmonary condition (r = - 0.68; 95CI: −0.82 to −0.46), or older adults (r = −0.40; 95CI: −0.60 to −0.14). We found no evidence of an association between kinesiophobia and physical activity in studies that included participants with chronic pain (r = −0.07; 95CI: −0.16 to 0.01) or acute pain (r = −0.13; 95CI: −0.45 to 0.23). Statistical heterogeneity was higher in the studies comprising people with a pulmonary condition (I^2^ = 98.1%), arthritis (I^2^ = 93.4%), or older adults (I^2^ = 91.2%) than in the studies comprising people with a cardiac (I^2^ = 28.7%) or neurologic condition (I^2^ = 55.9%).

The test of subgroup differences between self-reported (k = 54) and device-based (k = 29) measures of physical activity showed no evidence of a moderating effect of the type of physical activity measure (p = 0.171; Table 2). Both self-reported measures (r = −0.22; 95CI: −0.29 to −0.14; I^2^ = 89.3%) and device-based measures (r = −0.13; 95CI: −0.24 to −0.02; I^2^ = 57.2%) (Supplemental Figure 4) showed a negative association between kinesiophobia and physical activity.

We also found no evidence of a moderating effect of physical activity instruments (p = 0.209) (Supplemental Figure 5), physical activity outcome (p = 0.685) (Supplemental Figure 6), or kinesiophobia instrument (p = 0.452) (Supplemental Figure 7).

#### Meta-Regressions

Age did not statistically influence the correlation estimates of the meta-analysis studies (k = 72; p = 0.349). Similarly, the proportion of women (k = 72; p = 0.555) and the mean level of pain in the studies (k = 49; p = 0.481) did not influence correlation estimates.

#### Sensitivity Analysis

The meta-regression by quality score showed that a study’s quality did not influence correlation estimates (k = 83; p = 0.373).

## DISCUSSION

The main objective of this study was to systematically review and meta-analyze the direct relationship between kinesiophobia and physical activity. In addition, we examined the influence of potential moderators, such as health status. To our knowledge, this is the first review of its kind on this research topic.

### Kinesiophobia and Physical Activity

Both the main meta-analysis based on Pearson’s r correlation estimates and the secondary meta-analysis based on Spearman’s rho correlation estimates showed a small-to-moderate negative correlation between kinesiophobia and physical activity. Importantly, this correlation was observed bot in studies using both self-report (e.g., IPAQ) and device-based measures (i.e., accelerometers or pedometers). These results are consistent with our hypothesis and the dual models of physical activity^32–35^. According to these theoretical models, our findings suggest that the fear of movement characteristic of kinesiophobia triggers an impulse to avoid physical activity behaviors, which contributes to the maintenance or exacerbation of the initial fear. Accordingly, kinesiophobia and physical inactivity would be self-perpetuating or even self-reinforcing.

### Health Status

Our results suggest that patients with a cardiac, neurologic, arthritic, or pulmonary condition, as well as older adults, may be at greater risk for this negative relationship between kinesiophobia and physical activity than those with other conditions. In individuals with a cardiac condition, kinesiophobia and its impact on physical activity may be explained by a fear of inducing a new cardiac event^186^ or “causing more damage to the heart”^187^, but also by breathlessness (i.e., dyspnea), which reduces the ability to be physically active and damages confidence, leading to persistent anticipation of negative outcomes from physical activity^188^. Dyspnea is also a major barrier to physical activity in people with a pulmonary condition, such as chronic obstructive pulmonary disease (COPD)^147^. Patients with asthma may have additional disease-related barriers to physical activity, such as the fear of provoking respiratory symptoms and exacerbations^189^. Regarding neurologic conditions, chest tightness reported in patients with Parkinson’s disease as a barrier to exercise may be a factor contributing to the association between kinesiophobia and physical activity^190^. Another potential factor in these patients^190^, as well as in stroke survivors^191^ and healthy older adults^192^, is fear of falling. In patients with osteoarthritis, the belief that physical activity will “damage the joints”^193^ and the perceived fragility of their physical status^194^ may be factors contributing to the relationship between kinesiophobia and physical activity.

Although our results showed no evidence of an association between kinesiophobia and physical activity in other health conditions such as cancer, post-surgery, post-partum, or obstructive sleep apnea, these effects cannot be fully ruled out, as the lack of statistical significance could be explained by a lack of statistical power in these subgroup meta-analyses including fewer estimates (k = 1 to 5).

### Pain versus Fear

Our results showed no evidence of an association between kinesiophobia and physical activity in people with fibromyalgia, acute pain, or chronic pain. This finding was surprising because fear of pain is a key component of kinesiophobia, appearing in 10 of the 17 items on the TSK-17 and TSK-Heart scales, and reinforces the importance of considering the multidimensional nature of kinesiophobia, which not only relates to pain but also reflects fear of injury and fear of worsening a health condition.

In addition, contrary to our expectations, we found no statistical evidence showing that pain intensity at rest influenced the effect of kinesiophobia on physical activity, despite the substantial number of estimates included in this analysis (k = 23). This result is consistent with the weak relationship that has been shown between kinesiophobia and pain^195^, further suggesting that it is not the actual pain that prevents physical activity, but the fear of triggering pain, injury, or aggravating an underlying condition. However, this absence of evidence might be related to the methods used to assess pain, which may be better assessed by pain history (e.g., pain duration in months) or pain intensity during exercise.

### Limitations

The results of this systematic review and meta-analysis should be interpreted with consideration of several limitations. (1) We report considerable heterogeneity across the included studies, which may be explained by the diversity of the methods used to assess physical activity (questionnaires vs. accelerometers vs. pedometers), the instruments used in these methods (14 different questionnaires, 14 different accelerometers and pedometers), and the physical activity outcomes (n = 6), but also by the different questionnaires used to assess kinesiophobia (n = 11). This heterogeneity suggests that the measures of kinesiophobia and physical activity used in the literature reflect different dimensions of these two constructs. For example, self-reported measures of physical activity do not accurately reflect actual levels of physical activity^196^. (2) Because kinesiophobia is a state, i.e., a dynamic psychological variable, the time difference between the physical activity and kinesiophobia assessments, as well as the context of assessment, may have influenced the results. (3) While a subgroup meta-analysis showed no evidence of an effect of the type of TSK scale, inconsistencies have been noted in the purported dimensions assessed by different TSK scales or across populations^197^, which may have influenced our results. (4) Only 21 of the 98 authors we contacted (21%) shared their estimates (n = 13) or raw data (n = 8) with us, which is more than reported in previous literature^198^. Including these missing data may have affected the results.

## Conclusion

Higher levels of kinesiophobia were associated with lower levels of physical activity, especially in people with a cardiac, neurologic, arthritic, and pulmonary condition. According to theoretical models, this relationship between kinesiophobia and physical activity results from automatic processes that may be self-reinforcing and should therefore not be overlooked. However, heterogeneity between studies was substantial-to-considerable for some results, and the evidence for publication bias calls for cautious conclusions about this potential relationship. More evidence is required to determine the impact kinesiophobia should have on therapeutic decisions when aiming to maintain or increase physical activity. Particularly, prospective studies are needed to better understand the factors and mechanisms that influence the relationship between kinesiophobia and physical activity.

## Data Availability

All data, scripts, and materials used in the present study are available online at https://doi.org/10.5281/zenodo.11638244 (version 1.2).
The study was pre-registered in PROSPERO:
https://www.crd.york.ac.uk/prospero/display_record.php?ID=CRD42022364063

## ARTICLE INFORMATION

### Data and Code Sharing

According to good research practices^44^, the dataset, R Markdown script, and supplemental material are freely available in Zenodo^199^. A preprint version of this manuscript is publicly available online^200^ and has been recommended by Peer Community In Health & Movement Sciences^201^.

## Acknowledgements

Based on the Contributor Roles Taxonomy (CRediT)^202^ individual author contributions to this work are as follows: Miriam Goubran: Conceptualization, Methodology (Systematic Review), Investigation, Data Curation, Writing (Original Draft), Writing (Review and Editing); Ata Farajzadeh: Methodology (Systematic Review), Investigation, Data Curation, Writing (Original Draft), Writing (Review and Editing); Ian M. Lahart: Methodology (Meta-Analysis), Formal Analysis, Visualization, Data Curation, Writing (Original Draft), Writing (Review and Editing); Martin Bilodeau: Conceptualization, Methodology (Systematic Review), Writing (Review and Editing), Supervision (MG); Matthieu P. Boisgontier: Conceptualization, Methodology (Systematic Review and Meta Analysis), Investigation, Formal Analysis, Data Curation, Visualization; Writing (Original Draft), Writing (Review and Editing), Supervision (MG and AF), Project Administration, Funding Acquisition.

## SUPPLEMENTAL MATERIAL

**Supplemental Code 1.**
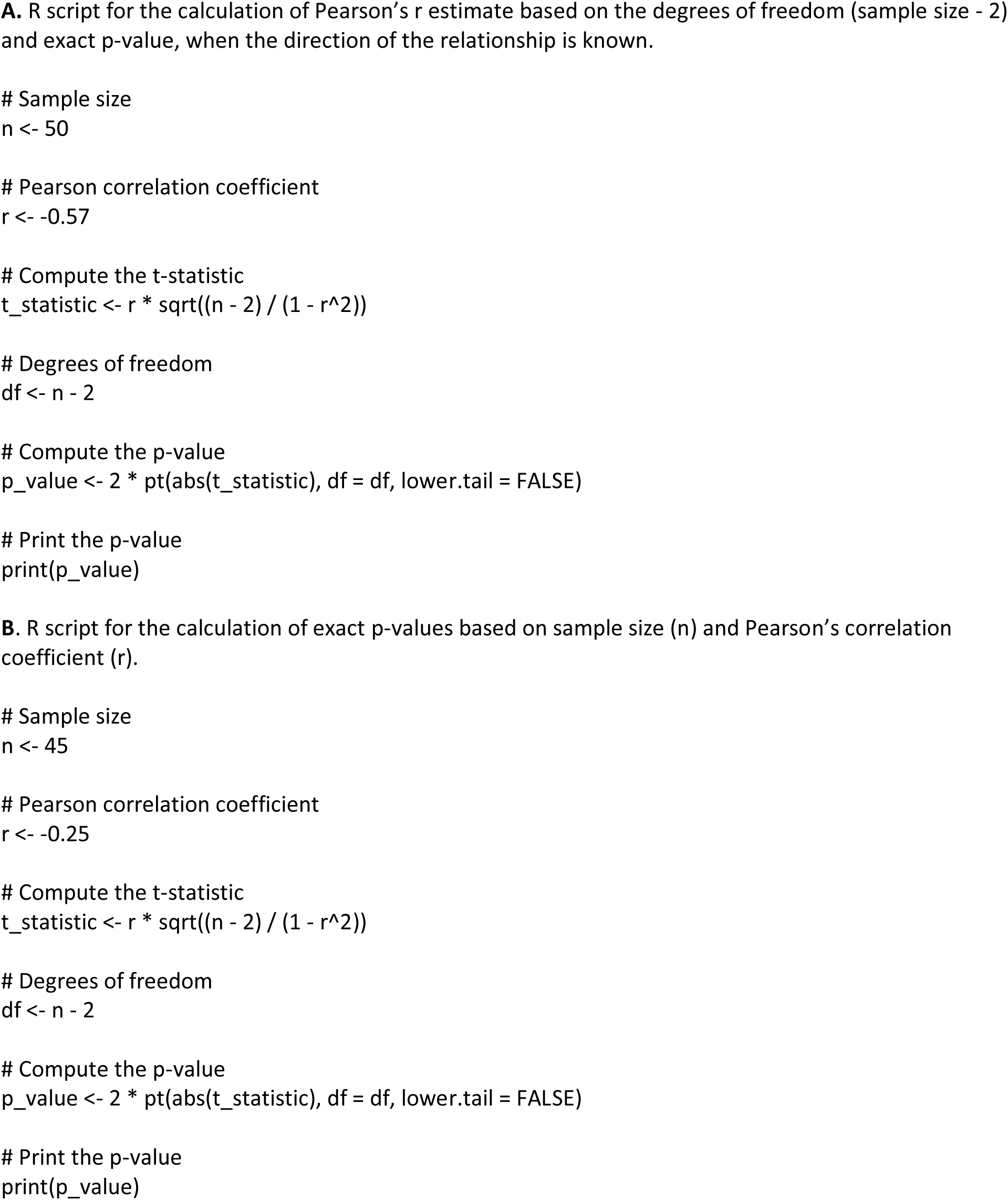
R scripts.

**Supplemental Figure 1.**
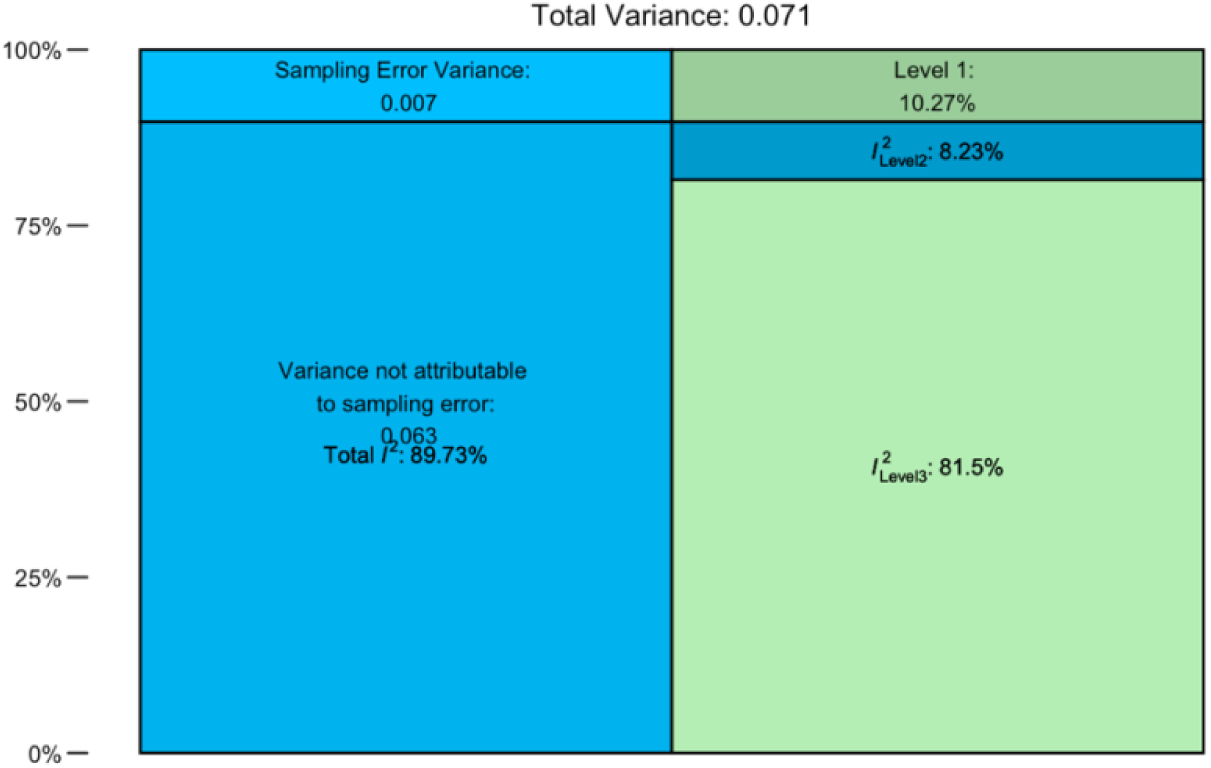
Heterogeneity variance.

**Supplemental Figure 2.**
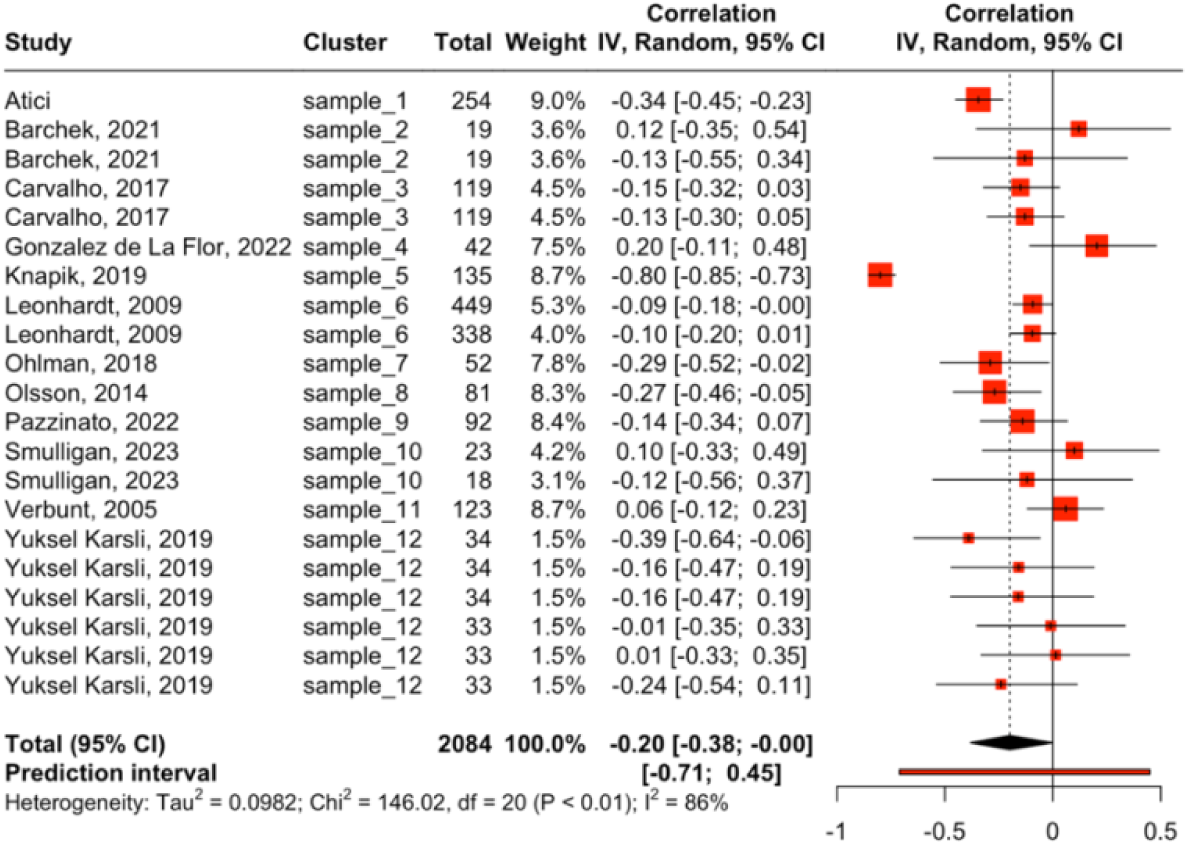
Secondary meta-analysis based on Pearson’s rho estimates.

**Supplemental Figure 3.**
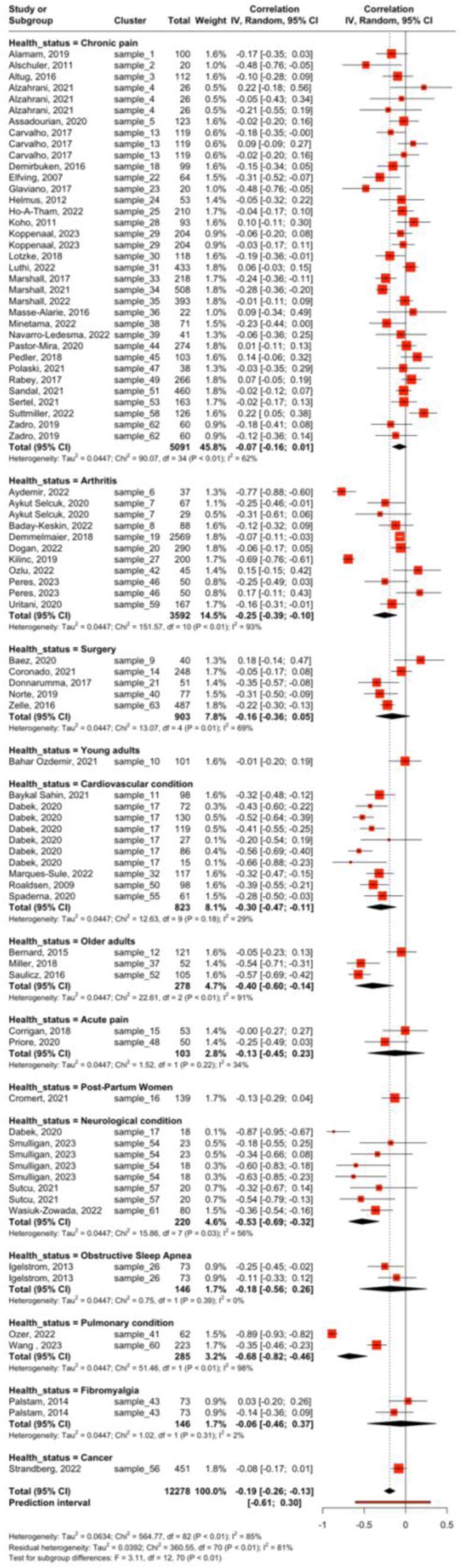
Subgroup meta-analysis: Differences by health status.

**Supplemental Figure 4.**
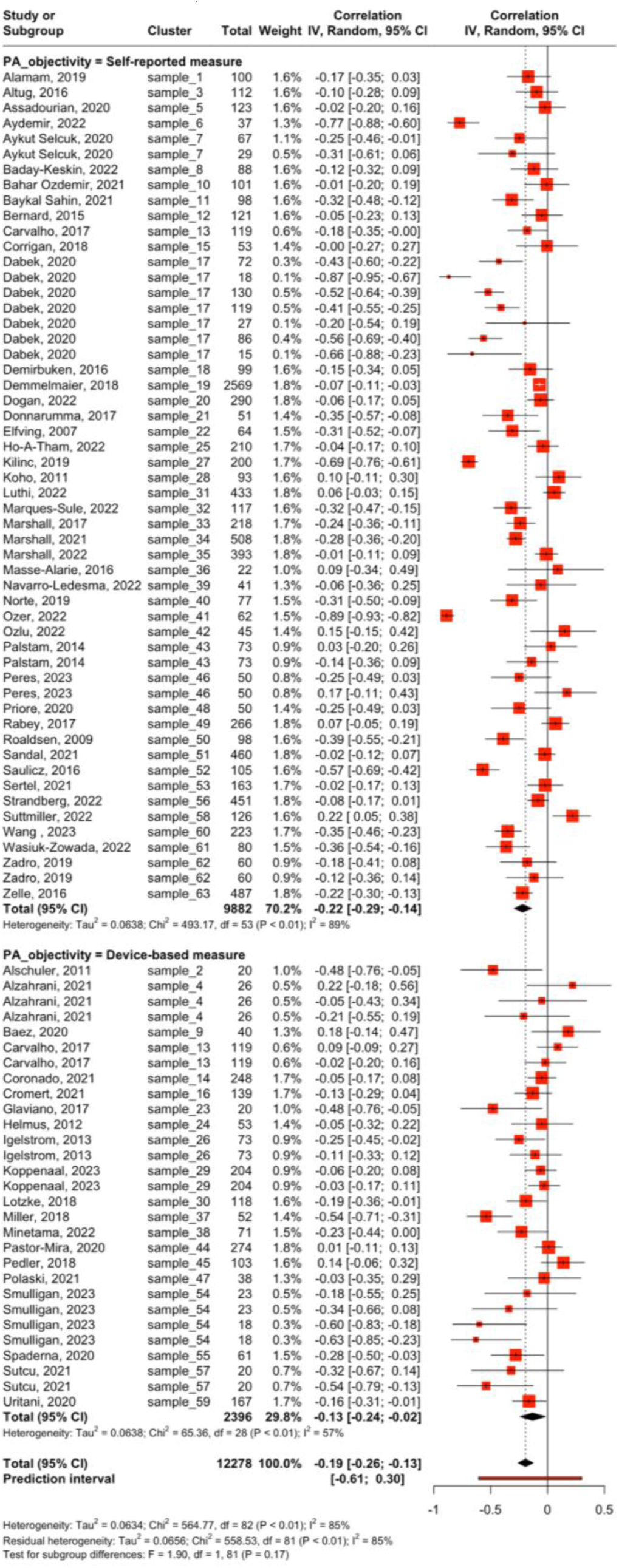
Subgroup meta-analysis: Differences by physical activity measure (self-reported vs. device-based).

**Supplemental Figure 5.**
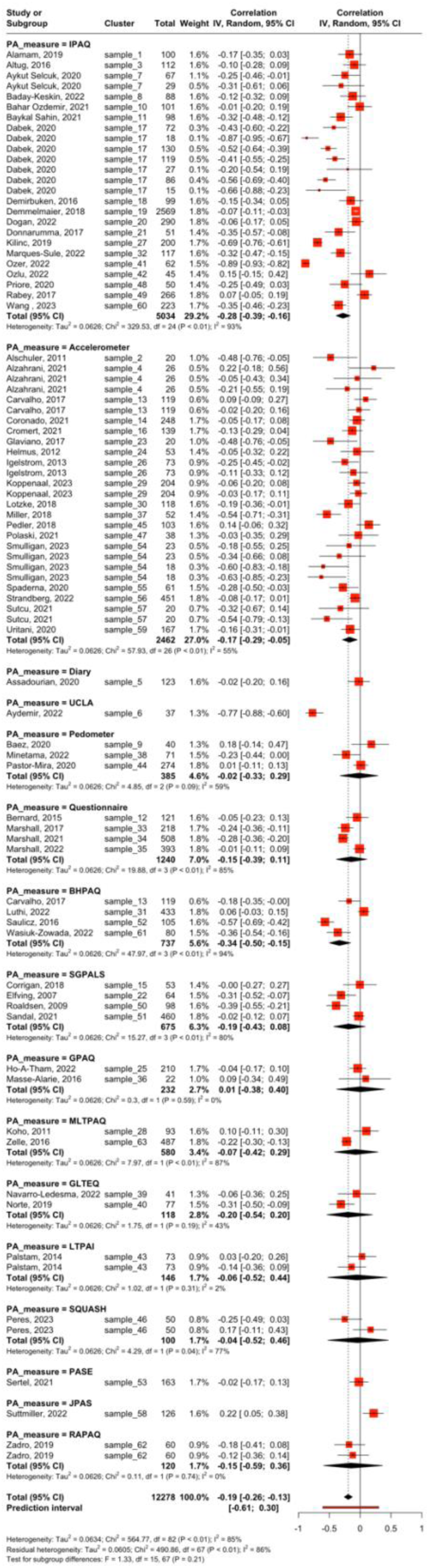
Subgroup meta-analysis: Differences by physical activity measurement instrument.

**Supplemental Figure 6.**
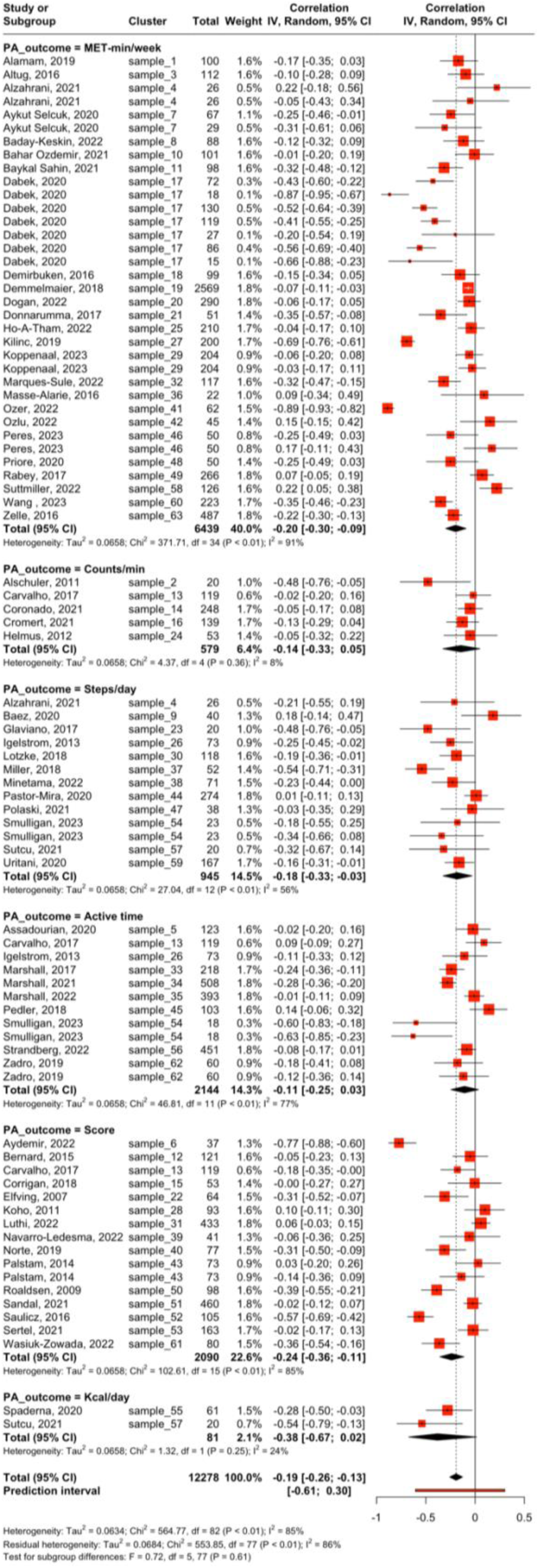
Subgroup meta-analysis: Differences by physical activity outcome

**Supplemental Figure 7.**
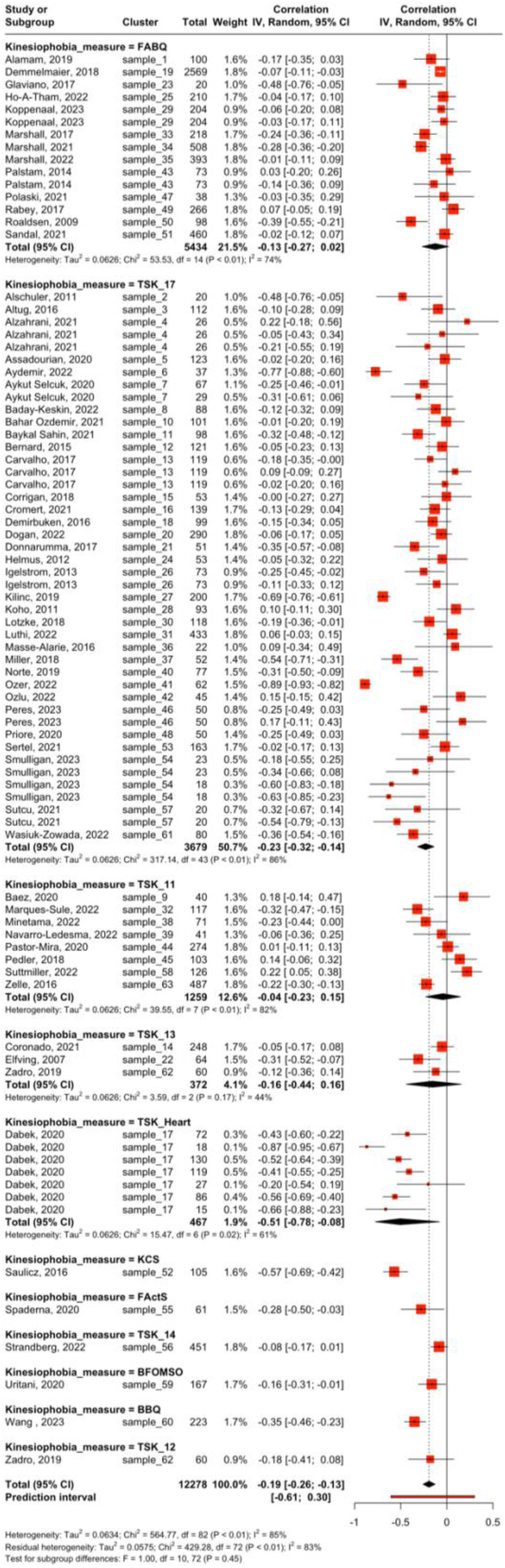
Subgroup meta-analysis: Differences by kinesiophobia measurement instrument.

